# Comprehensive Cerebral Aneurysm Rupture Prediction: From Clustering to Deep Learning

**DOI:** 10.1101/2024.10.31.24316531

**Authors:** Mostafa Zakeri, Amirhossein Atef, Mohammad Aziznia, Azadeh Jafari

**Affiliations:** School of Mechanical Engineering, College of Engineering, University of Tehran, P.O. Box 11155-4563, Tehran, Iran; Department of Mechanical Engineering, Virginia Tech, 24061, Virginia, USA

**Keywords:** Cerebral Aneurysm, Rupture Prediction, Machine Learning, Classification, Clustering, Recall

## Abstract

Cerebral aneurysm is a silent yet prevalent condition that affects a substantial portion of the global population. Aneurysms can develop due to various factors and present differently, necessitating diverse treatment approaches. Choosing the appropriate treatment upon diagnosis is paramount, as the severity of the disease dictates the course of action. The vulnerability of an aneurysm, particularly in the circle of Willis, is a critical concern; rupture can lead to irreversible consequences, including death. The primary objective of this study is to predict the rupture status of cerebral aneurysms using a comprehensive dataset that includes clinical, morphological, and hemodynamic data extracted from blood flow simulations of patients with actual vessels. Our goal is to provide valuable insights that can aid in treatment decision-making and potentially save the lives of future patients. Diagnosing and predicting the rupture status of aneurysms based solely on brain scans poses a significant challenge, often with limited accuracy, even for experienced physicians. However, harnessing statistical and machine learning (ML) techniques can enhance rupture prediction and treatment strategy selection. We employed a diverse set of supervised and unsupervised algorithms, training them on a database comprising over 700 cerebral aneurysms, which included 55 different parameters: 3 clinical, 35 morphological, and 17 hemodynamic features. Two of our models including stochastic gradient descent (SGD) and multi-layer perceptron (MLP) achieved a maximum area under the curve (AUC) of 0.86, a precision rate of 0.86, and a recall rate of 0.90 for prediction of cerebral aneurysm rupture. Given the sensitivity of the data and the critical nature of the condition, recall is a more vital parameter than accuracy and precision; our study achieved an acceptable recall score. Key features for rupture prediction included ellipticity index, low shear area ratio, and irregularity. Additionally, a one-dimensional CNN model predicted rupture status along a continuous spectrum, achieving 0.78 accuracy on the testing dataset, providing nuanced insights into rupture propensity.

## Introduction

Cerebral aneurysm is a silent brain disorder that affects between 2 and 5 percent of the world’s population^1^ and predominantly occurs in the proximal arterial bifurcations in the circle of Willis.^2^ Diagnostic methods for assessing aneurysms encompass a range of imaging modalities, including magnetic resonance (MR) and computed tomography (CT), each offering its own set of advantages and disadvantages.^3,4^

Formation, growth, and rupture of cerebral aneurysms can be attributed to various factors, including genetic and environmental factors,^5^ morphological and clinical factors,^6^ and hemodynamic factors.^7^ For example, a diameter-to-parent-vessel ratio greater than 2.3 (morphological factor),^8^ occurring in the anterior communicating artery (ACA) (clinical factor),^9^ containing complex unstable flow patterns^10^ and lower wall shear stress (WSS) (hemodynamic factor)^11^ are more susceptible to rupture. In a comprehensive study on the contribution of morphological features in rupture risk assessment, ellipticity index, size ratio, and irregularity were introduced as dominant features, respectively.^12^ Moreover, in a thorough study on the rupture prediction of cerebral aneurysms based on hemodynamic parameters, the low shear area ratio, relative residence time, and turnover time were recognized as the most important hemodynamic parameters, respectively.^13^

Given the small dimensions of the arteries within the circle of Willis, blood exhibits non-Newtonian behavior, and employing non-Newtonian rheology schemes can yield more realistic outcomes.^14,15^ Notably, the Carreau-Yasuda model^16^ is widely embraced for its ability to rectify the over-prediction of WSS by the Newtonian model and accurately represent the shear-thinning behavior at lower strain rates.^17^ This model aligns well with experimental data on blood behavior across different strain rates^18^ and demonstrates commendable performance in simulating pulsatile flows.^19^

Machine learning models have demonstrated the potential to outperform specialists in detecting cerebral aneurysms by leveraging vast datasets and advanced algorithms.^20^ Also, several studies have used ML to predict the rupture status of cerebral aneurysms. For instance, Tanioka et al.^21^ achieved a commendable accuracy of 0.78 by leveraging hemodynamic and morphological features based on data from 226 patients. In a separate investigation involving 615 patients, Random Forest (RF) models yielded an AUC of 0.81.^22^ However, cerebral aneurysm data are inherently sensitive, and metrics such as recall and precision are equally, if not more, crucial than accuracy.^23^ In this context, a study involving 124 patients achieved acceptable results with a precision of 0.80 and recall of 0.82 in predicting cerebral aneurysm rupture.^24^ In another study on aneurysm rupture risk estimation, k-nearest neighbor (KNN) achieved the highest recall score of 0.80, with a precision and AUC of 0.75 and 0.81, respectively.^25^ Furthermore, aggregating all available parameters appears to significantly enhance model performance, with accuracy gains ranging from 0.10 to 0.35 compared to considering only the five dominant parameters.^26^

This study aims to develop a rapid and precise method for predicting rupture in cerebral aneurysms. Our approach involves a comprehensive evaluation of 16 supervised and 10 unsupervised machine learning algorithms. In addition to assessing the models’ accuracy, we also emphasize evaluation precision and recall metrics, recognizing their increased sensitivity and importance in the context of this study. Moreover, we depart from the conventional use of the Newtonian model and instead employ the Carreau-Yasuda model to capture the shear-thinning behavior inherent in blood flow. This choice enables us to achieve a more accurate estimation of hemodynamic parameters distinguishing our work from most machine learning studies on cerebral aneurysms that predominantly relied on the Newtonian model. Furthermore, our study incorporates 55 clinical, morphological, and hemodynamic parameters. This comprehensive approach allows us to gain deeper insights into the dominant features contributing to aneurysm rupture and to compare the relative significance of each parameter in predicting rupture occurrence.

## Methodology

In this section, we present a comprehensive review of the database and study population. Following this, we briefly explain the parameters considered and the methods used for their extraction. Subsequently, we describe the data processing steps required to prepare the inputs for ML models and outline the ML algorithms used.

### Study Population

We employed 708 real geometries of cerebral aneurysms, including 253 ruptured and 455 unruptured cases extracted from patients in Sheffield, Milan, Geneva, and Barcelona, which were generously provided by the AneuX morphology database, an open-access, multi-centric database combining data from three European projects: AneuX project (www.aneux.ch), @neurIST project (www.aneurist.org), and Aneurisk (http://ecm2.mathcs.emory.edu/aneuriskweb/index).^27^ Further insights into the dataset are presented in Fig. 1, which provides valuable information regarding the distribution of aneurysm locations within the circle of Willis and the gender distribution of the patients. Notably, the Internal Cerebral Artery (ICA) emerges as the most common location for aneurysms, accounting for 323 out of the 708 cases considered. Next, the middle cerebral artery (MCA) and anterior communicating artery (ACA) are common locations for aneurysm formation in the circle of Willis in 181 and 143 cases, respectively. Moreover, we observe that the majority of the patients are females (520 females versus 188 males) with the number of female patients higher than male patients in each location.

**Fig. 1.**
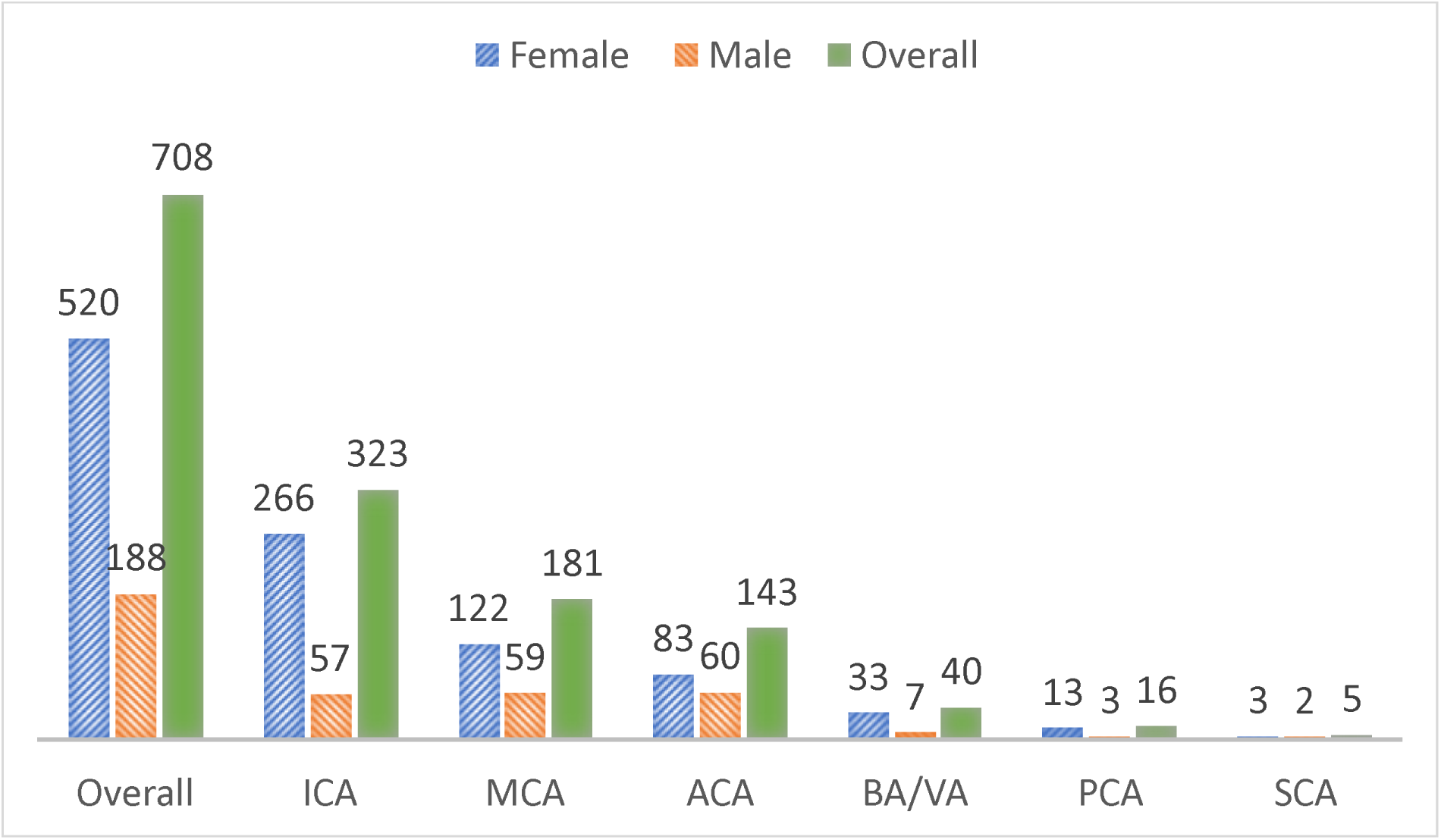
Scattering the dataset based on location and gender

To gain insights into the rupture status based on specific aneurysm locations and biological genders, we direct the reader to Fig. 2. This figure illustrates the rupture rates at different anatomical locations. According to the data presented in Fig. 2, the ACA exhibited the highest rupture rate, reaching 53.04% when considering both male and female patients combined. In contrast, the MCA had the lowest rupture rate (22.64% even though it was the second most common location for aneurysm formation). These findings shed light on the varying susceptibility of different aneurysm locations to rupture and provide valuable insights for our study.

**Fig. 2.**
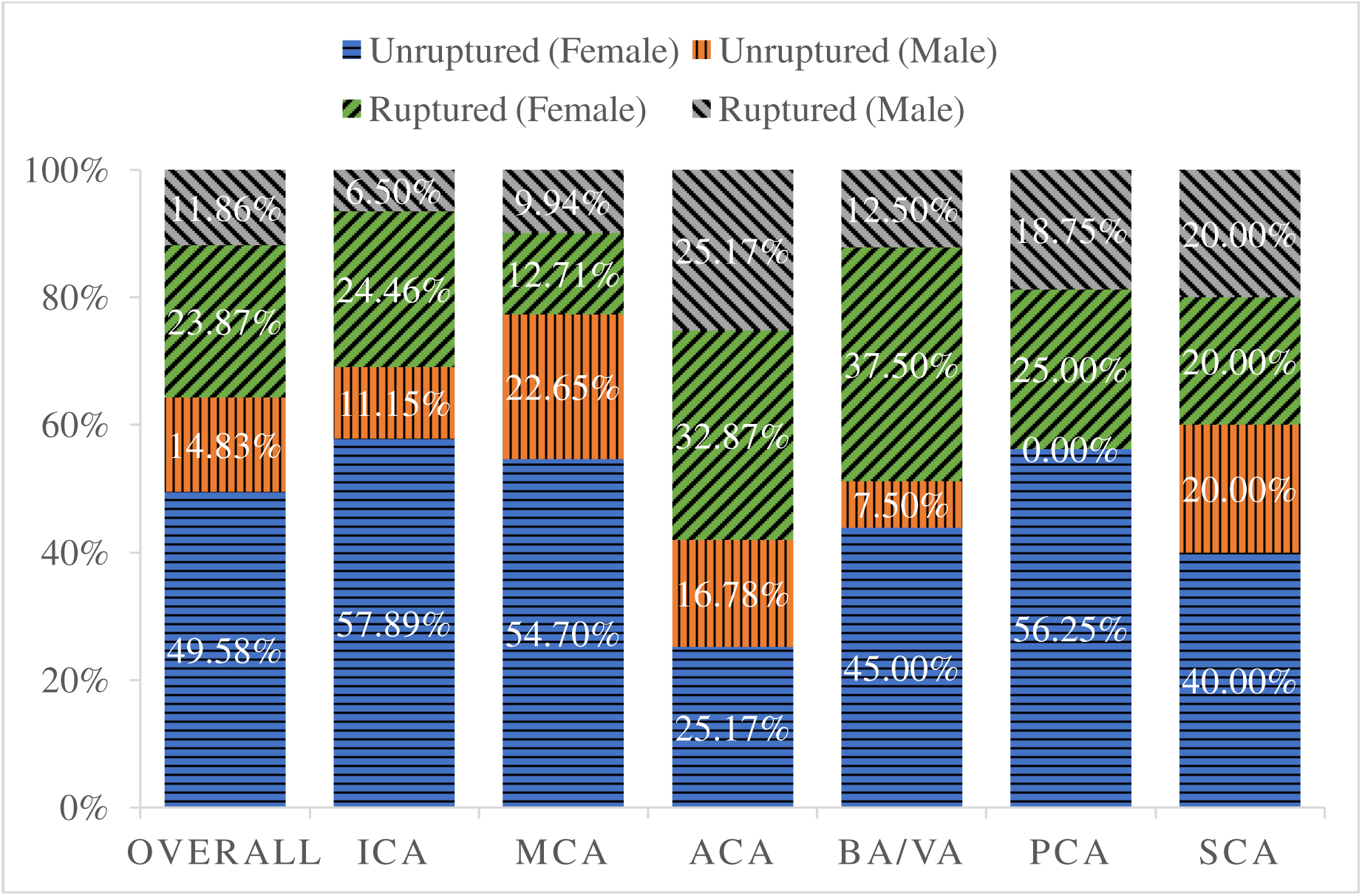
Rupture status scattering according to location and gender

### Blood Flow Simulation

In order to prepare the actual geometries obtained from medical imaging for simulation, it is necessary to complete a series of preprocessing stages. To address the imperfections inherent to the imaging process, such as rough surfaces and sharp corners resulting from errors and lack of accuracy, we employed the vascular modeling toolkit (VMTK) (http://www.vmtk.org/download/) and Materialize 3-matic v13.0. These tools facilitated the conversion of images to a suitable format for simulation tasks. We used COMSOL Multiphysics v6.0, employing the Carreau-Yasuda model to account for the shear-thinning behavior of blood. We considered a pulsatile velocity profile with scaling based on gender and aneurysm location. For more details about the simulation part, please refer to Zakeri et. al.^13^.

### Clinical, Morphological, and Hemodynamical Parameters

In total, our study encompasses 55 distinct parameters. Three of which are clinical variables: age, gender, and the location of the aneurysm within the circle of Willis. Additionally, there are 35 morphological features, including dome volume (DV), dome area (DA), maximum diameter (MD), central perpendicular height (CHP), neck diameter (ND), neck area (NA), aspect ratio (AR), bottleneck ratio (BR), diameter of the parent vessel at the inlet (DPVI), neck ratio (NR), size ratio (SR), maximum perpendicular height (MPH), irregularity (I), lateral or bifurcation status (LB), neck to dome ratio (NDR), volume to the ostium area (VOA), shaper factor (SF), in-flow angle (IMDA), aneurysm angle (MDNA), neck angle (INA), projection length (PL), ellipticity index (EI), non-sphericity shape index (NSI), undulation index (UI), maximum diameter parallel to the neck (MDPN), projection ratio (PR), conicity parameter (CP), neck circumference (NC), ideal roundness (IR), ideal roundness radius (IRR), ideal sphericity (IS), ideal sphericity radius (ISR), outflow number (ON), cumulative outlets diameter (COD), and inlet to outlet(s) ratio (IOR). Morphological features were extracted using COMSOL Multiphysics, Materialize 3-matic, and ParaView v5.11.0. Some of these results were derived from mathematical relations, while others were obtained directly from the original database. Additionally, 8 of these features were considered for the first time in our previous study. For further details regarding the definition of each parameter and the new 8 considered parameters, please refer to Zakeri et. al.^12^.

Furthermore, hemodynamic parameters were calculated through blood flow simulations conducted in COMSOL Multiphysics. The hemodynamic parameters were consistent with those previously reported by Zakeri et. al.^13^. The hemodynamics parameters included wall shear stress (WSS), normalized wall shear stress (NWSS), wall shear stress gradient (WSSG), wall shear stress divergence (WSSD), time-averaged wall shear stress (TAWSS), oscillatory shear index (OSI), aneurysm formation indicator (AFI), relative residence time (RRT), low shear area ratio (LSAR), oscillatory velocity index (OVI), flow velocity (FV), pressure loss coefficient (PLC), energy loss (EL), turnover time (TT), normalized pressure difference (NPD), flow stability (FS), and flow complexity (FC).

### Machine Learning

Before beginning the machine learning phase, it is advantageous to examine the data scattering (Fig. 3). This visualization effectively illustrates that linear models may not be suitable for our study. While Fig. 3 represents data scatter in three dimensions (LSAR, EI, and OVI), it is representative of the overall data distribution across all 55 dimensions in this study. Consequently, the complexity and nonlinearity of the data warrant the consideration of more sophisticated machine learning approaches.

**Fig. 3.**
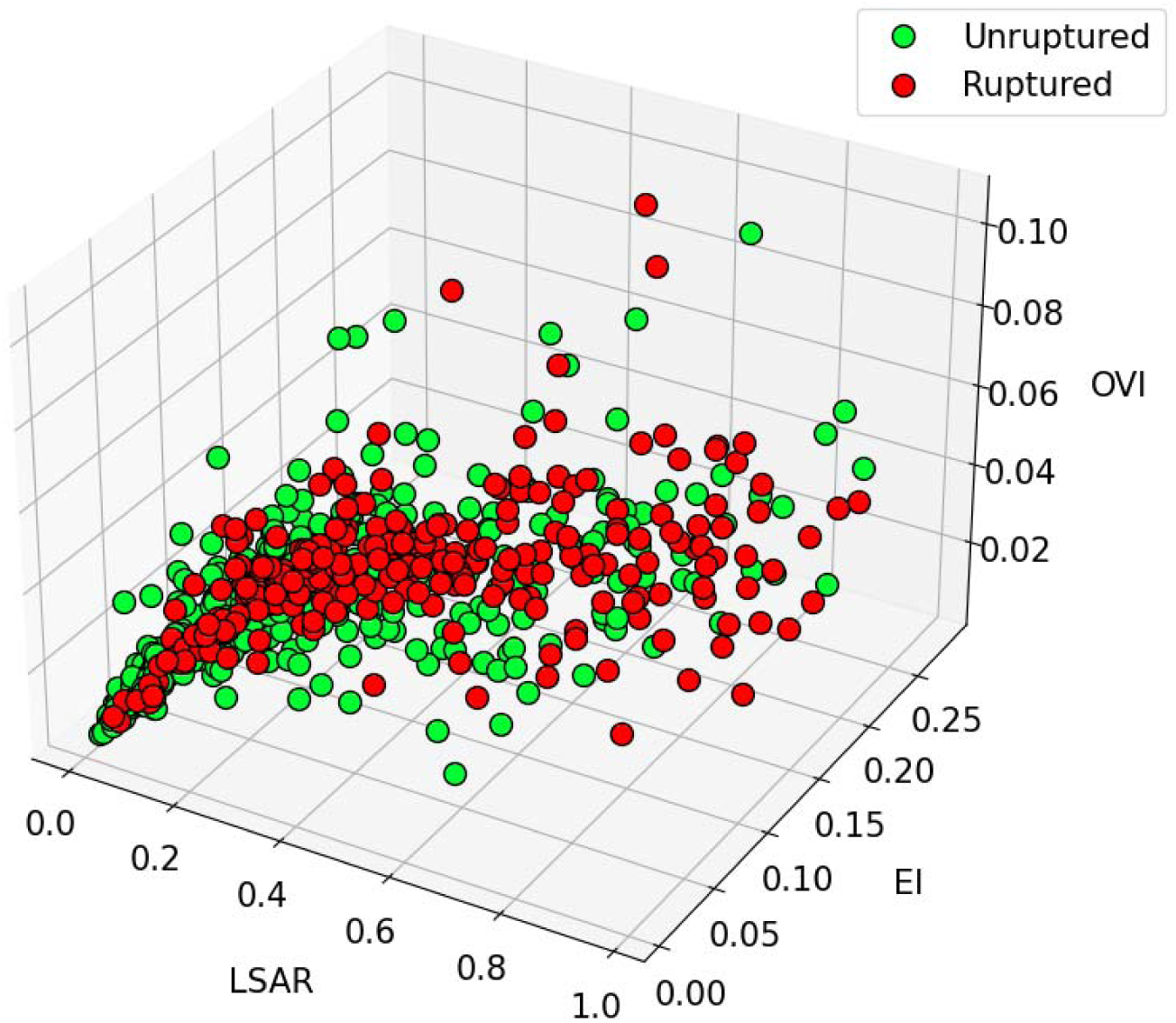
Scattering of data in a 3-dimensional space

after assembling the dataset, we employed standardization procedure to mitigate the influence of outliers. Subsequently, we used singular value decomposition (SVD) and principal component analysis (PCA) to identify the most influential features correlated with the rupture status of aneurysms. Nevertheless, after comparing the results of considering only the 10 dominant features recognized by PCA and all 55 features as inputs, we observed a notable enhancement in performance, with a minimal increase in training and prediction time when we considered the entire dataset as input.

In our comprehensive comparison of various machine learning algorithms, we trained 10 unsupervised models, 15 supervised models, and 1 semi-supervised model. This extensive approach allows us to address both classification and clustering concepts. The unsupervised algorithms employed include nearest neighbors, K-means, agglomerative, mean shift, affinity propagation, Birch, optics, spectral, Gaussian mixture, and DBSCAN. The aforementioned unsupervised algorithms are beneficial when dealing with datasets that lack any discernible rupture status for the patients. These algorithms are capable of identifying similarities between disparate features without prior knowledge of the rupture status, thereby clustering the features into two principal categories. Subsequently, the assistance of a specialist may be sought to ascertain the rupture status of the aneurysms in a subset of cases from each cluster. Since we were already aware of the rupture status of each case, we proceeded to identify each cluster based on its affinity with pre-existing labels related to rupture status. Finally, following the labeling of each cluster as either ruptured or unruptured, we used the accuracy definition to evaluate the performance of each unsupervised model. Additionally, the semi-supervised model, label spreading, was employed, which is advantageous when dealing with datasets that may contain missing labels.

To ensure robust and reliable results in the supervised evaluation, we implemented a 10-fold cross-validation. Subsequently, we combined the 10 models trained during the cross-validation process using a voting classifier with a soft-voting method. This approach yields robust models for each supervised model. Supervised algorithms encompass a wide array, including extreme gradient boosting (XGBoost), support vector machine (SVM), stochastic gradient descent (SGD), k-nearest neighbors (KNN), Gaussian process, Gaussian naïve Bayes, decision tree, and ensemble algorithms, such as bagging SVM, bagging KNN, random forest (RF), adaptive boosting (AdaBoost), gradient boosting, and histogram gradient boosting. Ensemble algorithms frequently modify or combine supervised models to enhance predictive performance.

In Addition, two additional supervised algorithms in the realm of neural networks were incorporated. These algorithms, multilayer perceptron (MLP) and sequential models, provide a higher degree of control over hyperparameters. In the training process of each model, we employed the grid search method to identify the most suitable hyperparameters for the given dataset.

## Results and Discussion

In this section, we begin by examining the outcomes of unsupervised models. We then conduct a comparative analysis of various supervised models, evaluating their performance in terms of accuracy, recall, and precision, and introduce the best models. Finally, we combine these top-performing models to create a superior model. To conclude, we discuss the significance of each parameter in predicting the rupture status of cerebral aneurysms.

### Clustering

It is important to recognize that unsupervised algorithms typically provide a narrow range of control than supervised algorithms. Consequently, it is often not possible to anticipate high-accuracy levels in clustering models. However, they are valuable in scenarios where labeled data are scarce or a significant portion of labels are missing. Fig. 4 illustrates the accuracy results of the clustering models trained on our dataset. Notably, the Birch and DBSCAN models obtained the highest accuracy (0.65). This signifies their ability to partition the data into two distinct groups with a 0.65 accuracy rate, which aligns with the actual labels in the dataset.

**Fig. 4.**
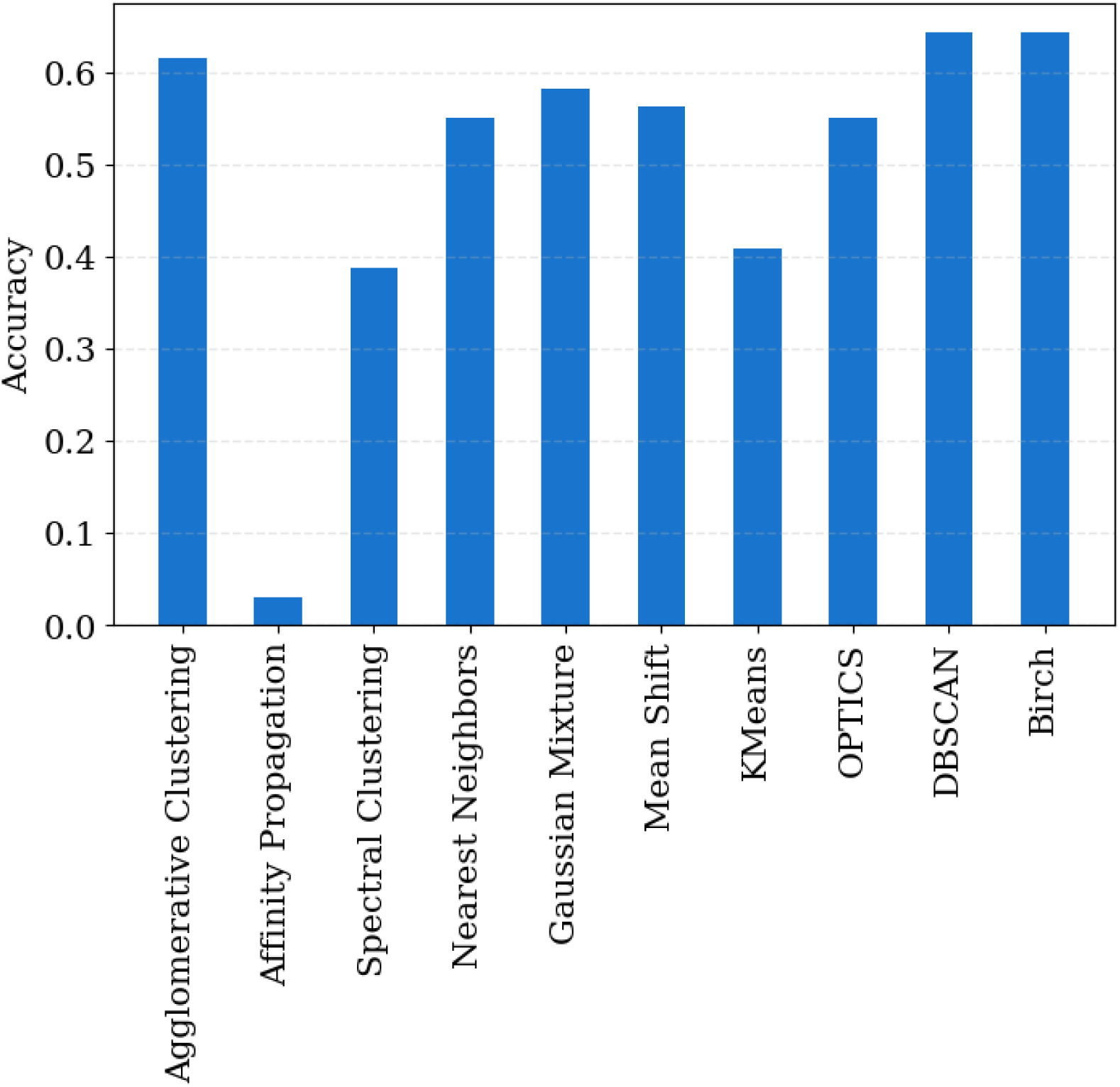
Clustering results

### Classification

In the field of machine learning, a prevalent methodology for evaluating the efficacy of a given model is to compare its accuracy on both training and testing datasets. The term “accuracy” is used to quantify the proportion of correctly predicted labels by the model. To ensure the model’s generalizability, it is advisable to avoid achieving an accuracy of 1 because this may indicate overfitting. Furthermore, it is crucial to maintain a minimal discrepancy (ideally no greater than 0.10) between the training and testing accuracy to guarantee the model’s capability to make accurate predictions on new, unseen data. As shown in Fig. 5, the SGD, SVM, and MLP models emerge as the top-performing classification models, exhibiting an accuracy range of 0.82 to 0.85 on the testing data. This demonstrates their commendable capacity to make accurate predictions of unseen data. Conversely, the Gaussian Naïve Bayes model exhibits the least favorable performance with an accuracy of 0.76 on the testing dataset. The semi-supervised model, label spreading, appears to be adversely affected by potential overfitting on the training data, reaching an accuracy of 1 on the training dataset and 0.76 on the testing dataset. Therefore, this model, along with histogradient boosting and self-training, is overfitted within the context of this study.

**Fig. 5.**
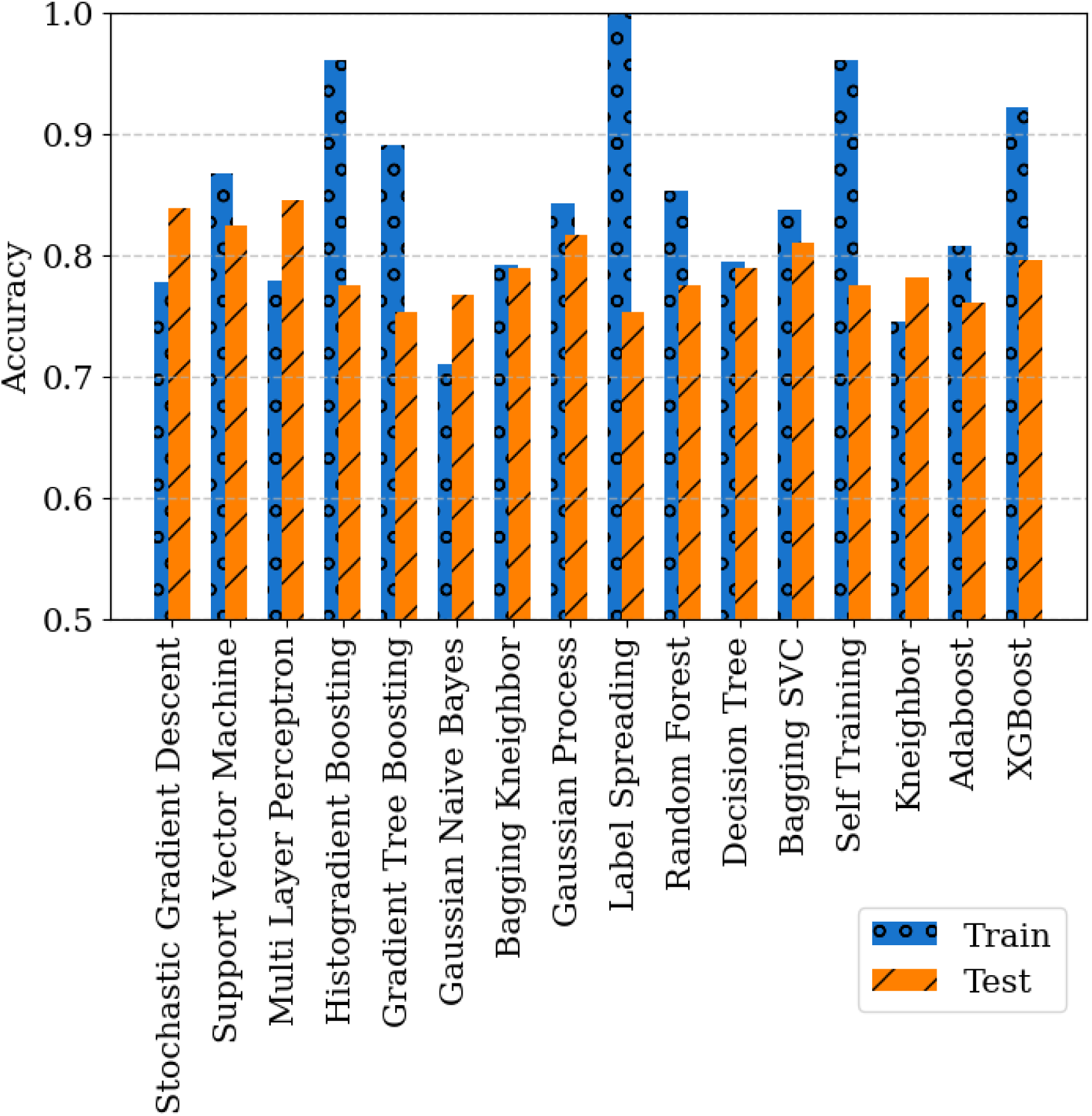
Classification accuracy of training and testing datasets

Given the medical nature of our dataset, it is important to underscore that alternative evaluation metrics, such as precision and recall, hold greater significance compared to accuracy. Recall can be defined as the model’s ability to correctly identify occurrences when it predicts their likelihood, whereas precision signifies the model’s ability to correctly predict most occurrences. Consequently, while recall is considered a pivotal factor, it is crucial not to disregard accuracy and precision because they contribute to the overall model performance. In this context, we considered all three factors: accuracy, precision, and recall. As illustrated in Fig. 6, the SGD, SVM, and MLP models once again demonstrate the highest performance. The precision of these models falls within the range of 0.83 to 0.87, while recall falls within the range of 0.90 to 0.92. It is noteworthy that although there are models with superior recall scores, their inferior performance in terms of precision and accuracy has led us to conclude that the SGD, SVM, and MLP models are the optimal choices due to their balanced performance across all three metrics.

**Fig. 6.**
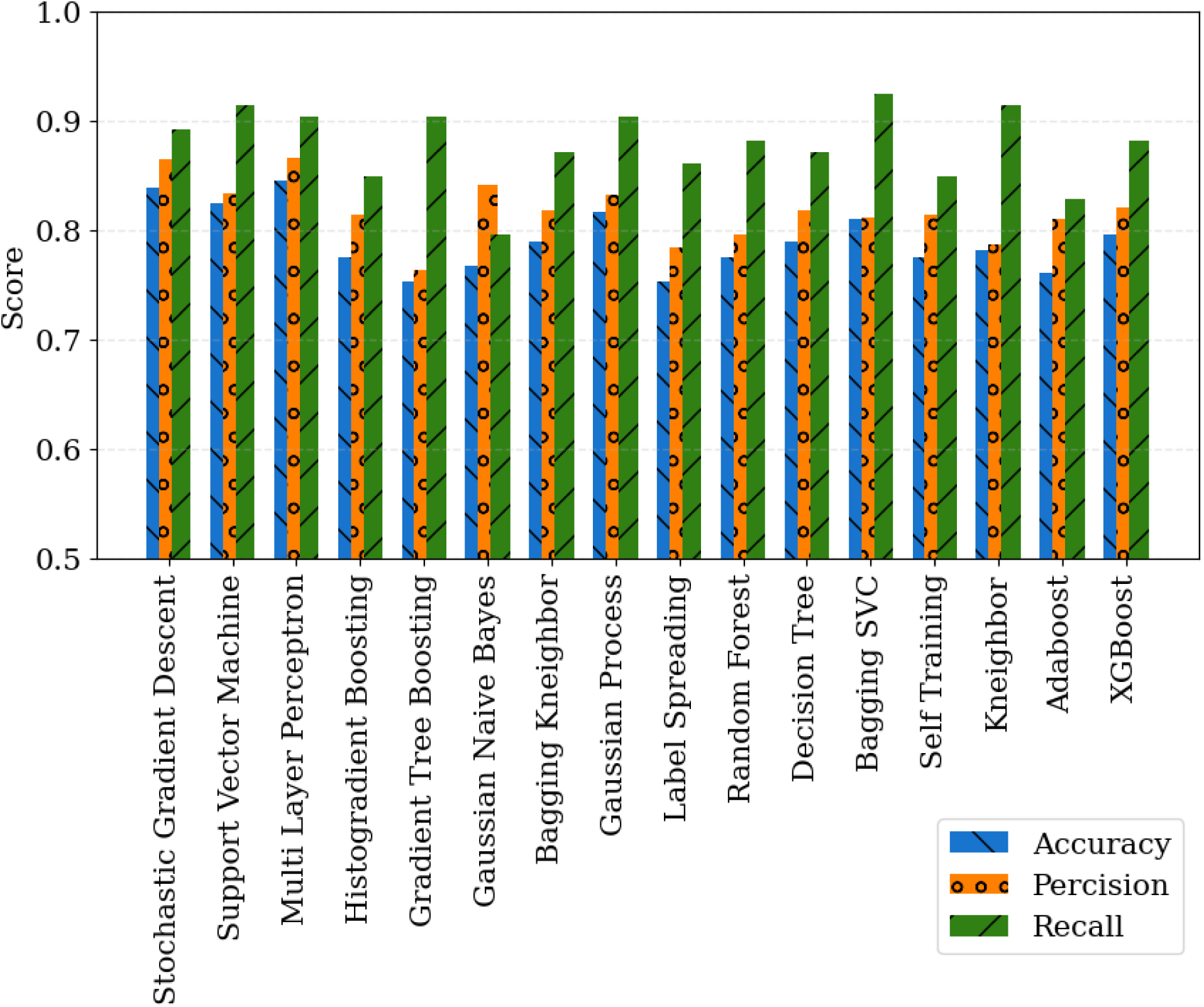
Accuracy, precision, and recall of the testing dataset

### ROC Curve

The receiver operating characteristic (ROC) curve visually represents the balance between true and false positive rates. In the case of the random classifier, the ROC curve is linear, indicating that the true and false positive rates are equal. As a model demonstrates enhanced performance, its ROC curve shifts towards the upper left corner. In an ideal scenario, the model should achieve a true positive rate of 1 and a false positive rate of 0. The area under the curve (AUC) serves as a crucial metric for evaluating model performance. An AUC of 0.5 indicates a random classifier, whereas an AUC of 1 represents a model with perfect classification capabilities. Fig. 7 illustrates the ROC curves and the corresponding AUC values for each model. In consideration of this metric, the SGD and MLP models exhibit notable performance, with ROC curves that follow favorable paths and AUC values of 0.86. The SVM model is positioned at the next level, with an AUC of 0.85, and it ties with other models like Gaussian process and bagging SVM. In contrast, the Gaussian Naïve Bayes model demonstrates comparatively diminished performance with an AUC of 0.78. All models achieved results that were either equal to or superior to this score.

**Fig. 7.**
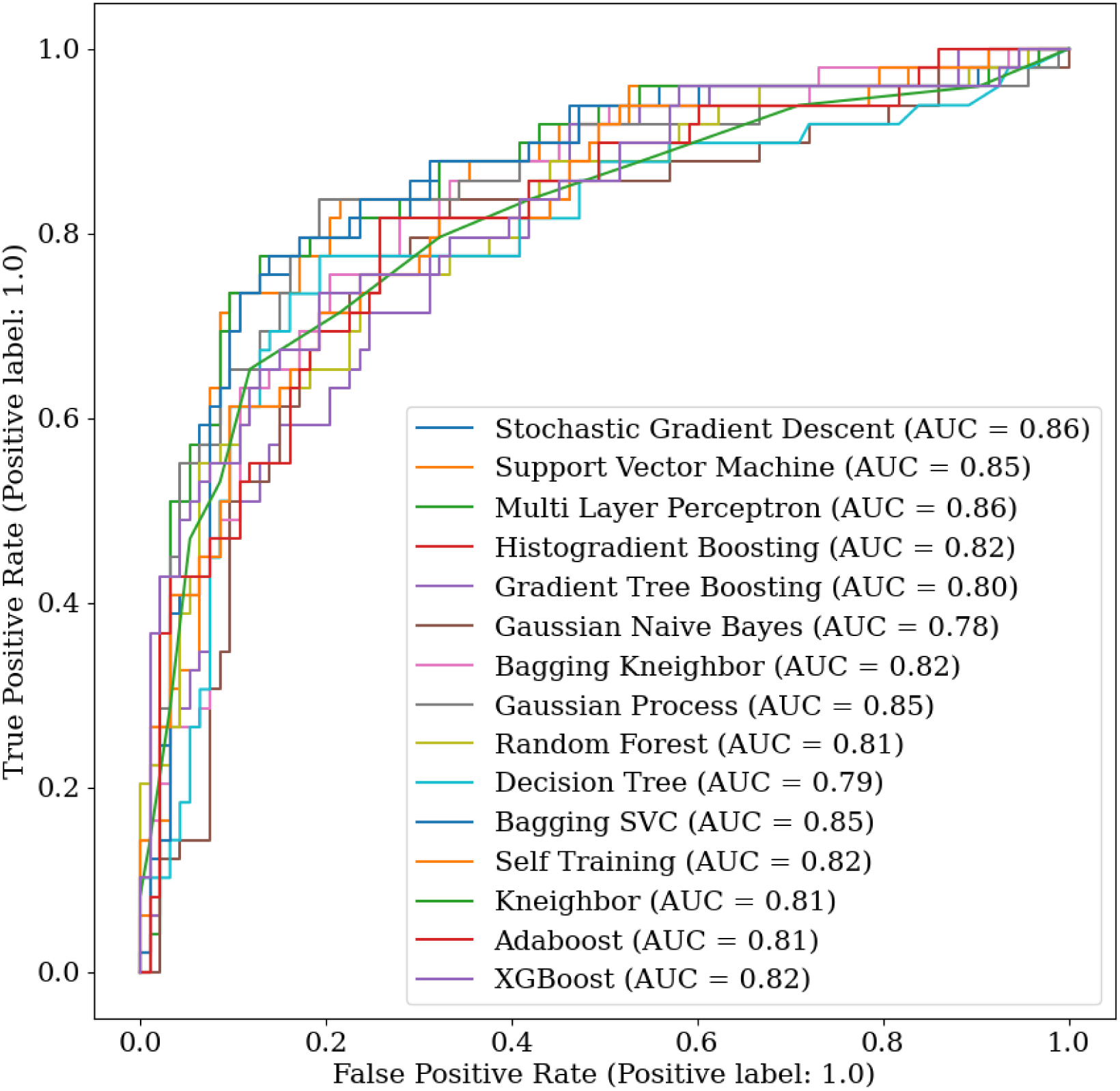
Receiver Operating Characteristic (ROC) Curve for all models

### Superior model by combining best models

Given that we typically employ shuffle splits for training and testing data, it is common to obtain disparate model outputs when training is conducted. Furthermore, each model has a unique perspective relative to data categorization. To capitalize on the strengths of multiple models, we employed a voting classifier model, which combines the three most effective models to create a superior model that incorporates the advantages of each. The model metrics were as follows: an accuracy of 0.83 for the training dataset, 0.84 for the testing dataset, with a precision of 0.86 and a recall of 0.90. The ensemble model produces results that are comparable to those of the best standalone models. However, its primary advantage lies in its enhanced stability across multiple training iterations. It is crucial to underscore that, in this context, where medical data are involved, recall and precision are of paramount importance. Therefore, with recall and precision scores of 0.90 and 0.86, respectively, this ensemble model demonstrates its efficacy in handling medical data’s specific requirements.

### Dominant Features

Each model employs a unique algorithm for data subsampling, resulting in disparate parameter values across models. Fig. 8 Feature importance for the three best permutation models illustrates the most significant factors using the permutation method, which assesses the correlation between input features and the model’s performance by measuring the extent to which the model’s prediction accuracy is compromised when the values of a particular feature are randomly shuffled. According to Fig. 8. ellipticity index (EI), low shear area ratio (LSAR), and irregularity (I) occupy the top positions of the three best models, respectively. Notably, the significance of these features is more pronounced in the MLP, SVM, and SGD models in that order. The first and third influential parameters are morphological in nature, whereas the second parameter, LSAR, is a hemodynamic parameter. Another hemodynamic feature, normalized wall shear stress (NWSS), emerges as the fourth parameter in terms of importance for SGD and SVM, while it is ranked as the fifth feature for MLP. In addition, projection length (PL) ranked fourth for MLP and fifth for SGD and SVM. Conversely, we observe some negative correlations between certain features and permutation results. This indicates that the model’s accuracy remains unaffected by the shuffling of data related to that feature, and in some cases, may even improve. In other words, the specific feature in question plays no significant role in the model, and data shuffling did not affect the model’s accuracy. In fact, it may have improved the model’s performance, indicating a lack of correlation between that feature and the model’s accuracy or performance. Nevertheless, it would be erroneous to conclude that this feature can be ignored or removed from the dataset, since it might be an important feature for other models. In addition, the correlation between the inputs and accuracy of the models may vary each time the test is run due to the randomization process. However, the most important features are stable after each randomization and training, and variations occur in the less important or unstable features. For example, the wall shear stress gradient (WSSG) is ranked as the 12th most important feature among all 55 features in the SVM model, while it is one of the worst features in the other two models.

**Fig. 8.**
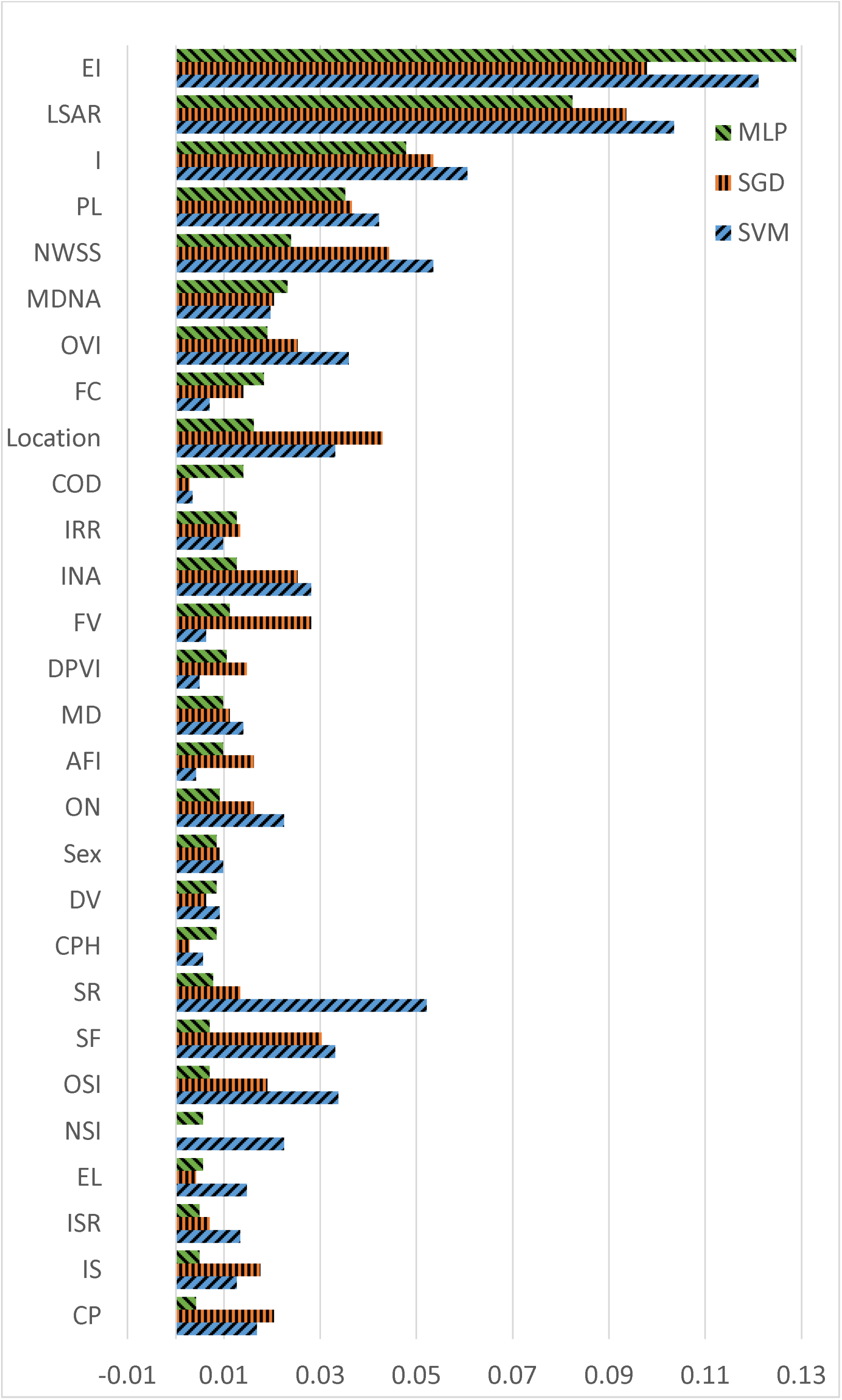
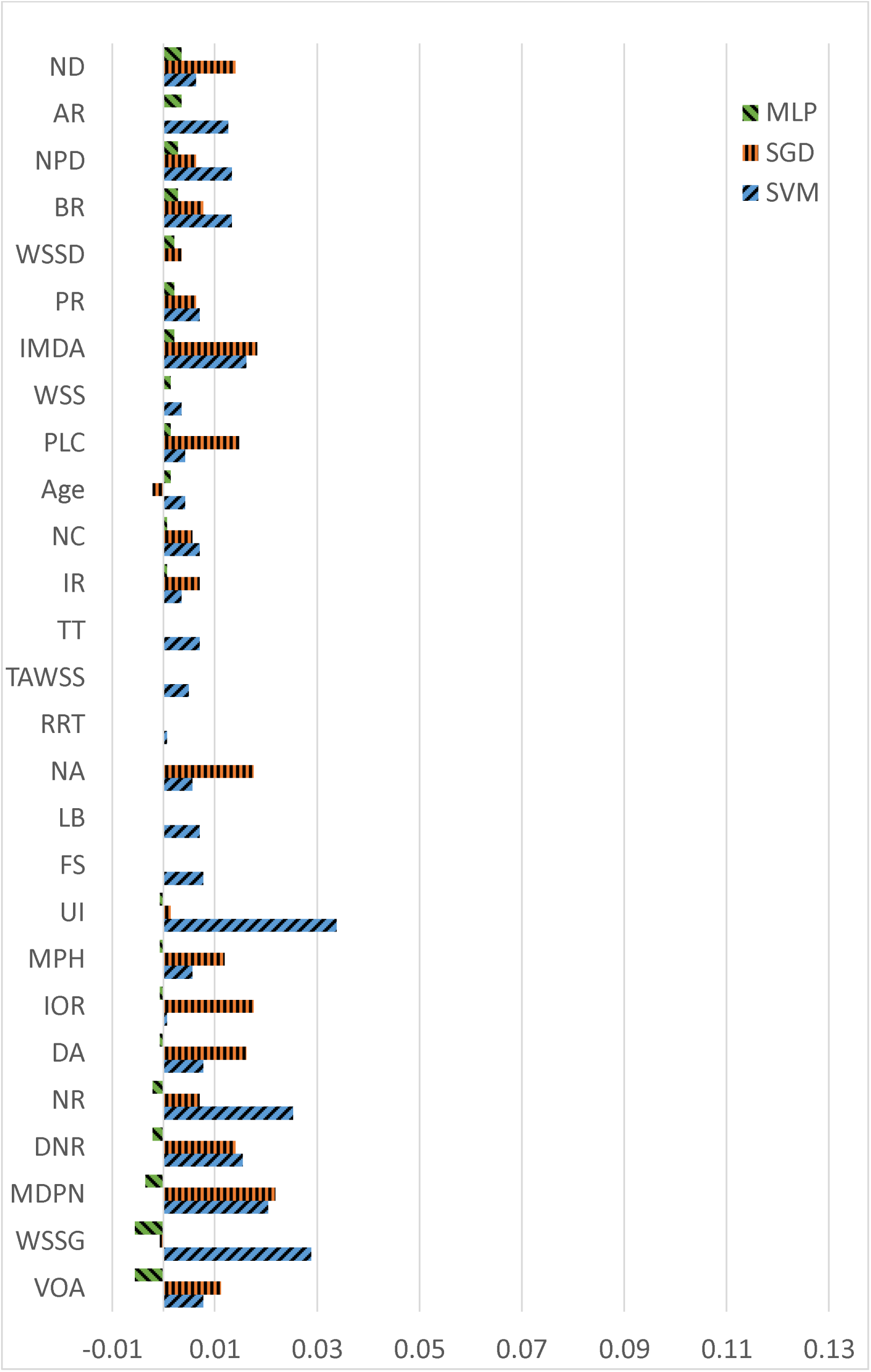
Feature importance for the three best permutation models

It is noteworthy that, despite relative residence time (RRT) being identified as a neutral or less important parameter in our study, it might be a key factor in other studies. In addition, lateral or bifurcation (LB) status may be regarded as a pivotal parameter by physicians, despite not being identified as a significant parameter in this study. The most plausible explanation for this inconsistency is the number of features included and the manner in which they are compared. To illustrate, LB may be a crucial parameter when considered in isolation; however, its importance may diminish when all parameters are considered. In this context, other parameters may be more effective in predicting rupture in cerebral aneurysms.

### Probabilistic Rupture Prediction

In the final phase of this study, we developed a one-dimensional convolutional neural network (CNN) model to predict aneurysm rupture status. The model architecture includes 1D CNN, max pooling, batch normalization, dropout, and dense layers, resulting in a robust model with an accuracy of 0,78 on the testing dataset. A key feature of this model is the use of a sigmoid activation function in the output layer, while all other layers employ ReLU activation. This configuration enables the model to predict rupture status along a continuous spectrum, in contrast to previous binary predictors. The model output is a real number between 0 and 1, with values below 0.5 classified as unruptured and those above 0.5 as ruptured. Importantly, within the ruptured category, higher prediction values indicate a greater likelihood of rupture and greater similarity to known ruptured cases, potentially correlating with the severity of the patient’s condition. Conversely, cases classified as unruptured but with values approaching 0.5 may signal a risk for potential future rupture, possibly due to aneurysm growth and development.

## Conclusion

We used a comprehensive dataset comprising over 700 authentic cerebral aneurysm geometries, encompassing 3 clinical, 35 morphological, and 17 hemodynamic features for each geometry. The primary objective of this study was to assess the individual contributions of these parameters to predicting cerebral aneurysm rupture status. In our analysis, we trained 16 classification models and 10 clustering models to predict aneurysm rupture status based on the selected parameters. To capture the shear-thinning behavior of blood in the small arteries of the circle of Willis, we employed the Carreau-Yasuda model for hemodynamic feature extraction. The clustering section achieved an accuracy of 0.65, with the Birch and DBSCAN models demonstrating the greatest efficacy. In the classification section, the SGD and MLP models exhibited the highest performance, achieving an AUC of 0.86, along with recall, and precision rates of approximately 0.90 and 0.86, respectively. To enhance the stability of our results, we combined the three most promising classification models (SGD, MLP, and SVM) into a superior ensemble model using a voting classifier, ensuring robustness against data variations. Among the identified parameters, certain factors showed significant correlation with the performance of the best models. These include the ellipticity index (EI), low shear area ratio (LSAR), and irregularity (I), which emerged as the most dominant features for predicting rupture status. Following these, the normalized wall shear stress (NWSS) was identified as the fourth most significant feature in the SGD and SVM models and the fifth in the MLP. Additionally, the projection length (PL) ranked fourth in the MLP and fifth in the SGD and SVM models. These important features can be classified into two categories: morphological parameters (EI, I, and PL) and hemodynamic parameters (LSAR and NWSS). Finally, the implementation of a 1D CNN model with continuous output demonstrates potential for more nuanced risk assessment in aneurysm cases, offering valuable insights for clinical decision-making and patient monitoring.

## Data Availability

The data that support the findings of this study are available from the corresponding author up-on reasonable request.

## Funding

No funding was received towards this work.

## Competing interests

The authors report no competing interests.

## Notes

### Competing Interest Statement

The authors have declared no competing interest.

### Funding Statement

No external funding was received.

### Summary of Updates

One of the reference has been updated

